# The causal correlation between gastroesophageal reflux disease and chronic widespread pain: a bidirectional mendelian randomization study

**DOI:** 10.1101/2024.05.06.24306927

**Authors:** Menglin Chen, Houshu Tu, Jiaoli Zhou, Yi Zhang, Shuting Wen, Yao Xiao, Ling He

## Abstract

**Background:** Previous observational research found a relationship between gastroesophageal reflux disease (GERD) and chronic widespread pain (CWP). Despite this, it is unknown which, if any, of the conditions produces the other. Our study will use bidirectional Mendelian randomization (MR) to evaluate their causal link.

**Methods:** We examined two sets of publically accessible data from genome-wide association studies (GWAS): GERD (129,080 cases and 602,604 controls) and CWP (6,914 cases and 242,929 controls). We used the inverse variance weighting (IVW) approach as the major analysis method, but we also ran weighted median and MR-Egger regression analyses. We performed various sensitivity studies to assess the conclusions’ consistency, horizontal pleiotropy, and stability.

**Results:** MR analysis showed that CWP increased the risk of developing GERD [N_SNP_ = 4, odds ratio (OR): 245.244; 95% confidence interval (CI): 4.35E+00,1.38E+04; p = 0.007 < 0.05] and vice versa (N_SNP_ = 28; OR:1.019; 95% CI: 1.009-1.029; p = 0.029 < 0.05). Bidirectional evidence of causality existed. The sensitivity analysis demonstrated the robustness and reliability of the findings.

**Conclusions:** Our study demonstrated a bidirectional causal relationship between GERD and chronic widespread pain, and future interventions for CWP may be an effective strategy for preventing or mitigating GERD and vice versa.

## Introduction

Gastroesophageal reflux disease (GERD), which causes heartburn and regurgitation, is a prevalent chronic gastrointestinal condition (1). It impacts up to 20% of the population in the West, and its incidence is rising globally as the world’s population grows and ages (2, 3), posing a major public health concern.

Chronic widespread pain (CWP) is clinically defined as musculoskeletal discomfort with a diffuse pattern that lasts three months or more (4). It is one of the characteristic symptoms of fibromyalgia (FM) condition, involving 10.6% to 11.8% of the general population (5). Meanwhile, strong research indicates that patients who suffer from CWP have an increased chance of mortality (6). This emphasizes the importance of proactive risk-reduction s initiatives, as well as early prevention and treatment for illness. Prioritizing early detection, diagnosis, and management of CWP is critical for improving patients’ overall health outcomes.

It is commonly acknowledged that the term FM is the most precise approach to characterize CWP after all other plausible explanations have been ruled out (4). Retrospective research discovered that patients with FM commonly have GERD as a secondary disorder (7). Meanwhile, previous cross-sectional research discovered a positive link between FM and GERD (8), signaling that persons with CWP may be more likely to acquire GERD. Currently, research suggests that CWP is linked to a variety of illnesses, including anxiety, depression, sleep difficulties, and migraine headaches (9). However, there is no clear evidence that CWP causes GERD. Mendelian randomization (MR) is a great approach for researchers who want to avoid confounding biases and causal inversion (10, 11). To ensure the validity and reliability of its results, it uses genetic variants, namely single nucleotide polymorphisms, or SNPs, as instrumental variants (IVs) (12).

The aim of the research is to evaluate the causal relationship between GERD and CWP via a two-sample bidirectional MR analysis (MRA) method, which will provide some basis for etiology and treatment.

## Methods

The present study exploited genetic information from an extensive publicly accessible database of genome-wide association studies (GWAS), which required no further ethical approval.

### Research design

This study employed two-sample MRA to determine the causal link between CWP and GERD. This study tested the following primary hypotheses (13): (1) SNPs were significantly related with exposure. (2) The SNPs showed no correlation with putative confounders of the CWP-GERD relationship. (3) SNPs altered the result solely by connection with the exposure. This work was designed and written in accordance with the STROBE-MR reporting criteria (14). Fig 1 displays the general process chart.

**Fig 1.**
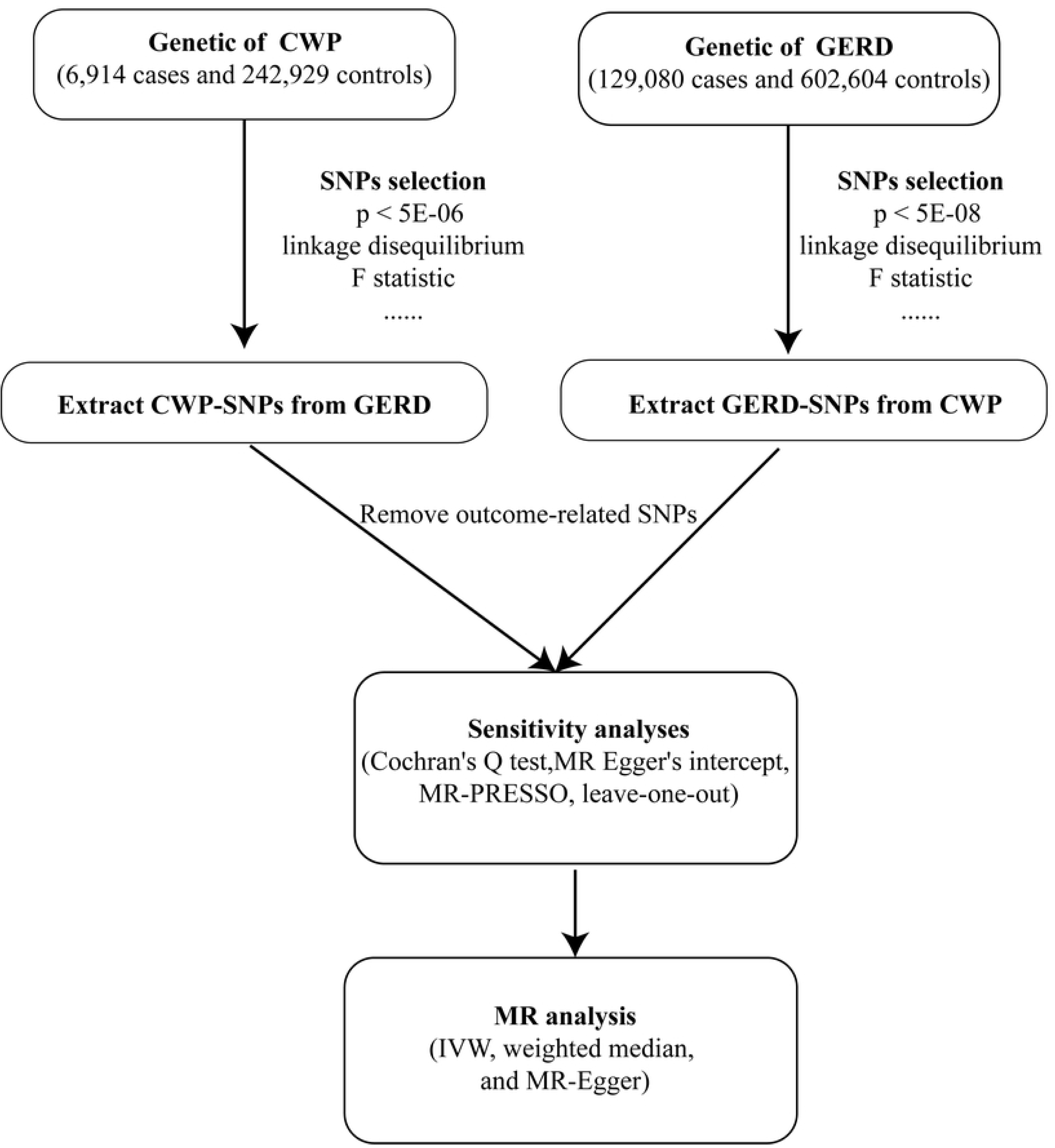
The general process chart of the MR study.

### Data sources

A large-scale GWAS comprising 473,524 controls and 129,080 patients provided the GERD data (15). The CWP data, on the other hand, came from a GWAS investigation at the UK Biobank that included 6914 cases and 242,929 controls (16). The patients were categorized based on the self-identified diagnosis of fibromyalgia and/or the presence of more than three months’ worth of knee, arm, hip, spine, or generalized discomfort. Exclusion criteria included people with systemic lupus erythematosus, ankylosing spondylitis, arthritis of the joints, rheumatic polymyalgia, and myopathy.

It is important to emphasize that there was no sample duplication between the two research groups and that every participant was of European heritage, reducing the possibility of ethnic bias. The study met with ethical standards and legal requirements and used data that was already publicly available. As a result, no further ethical assessment or clearance was needed.

### Screening of SNPs

In accordance with the MR hypothesis criterion, an SNP correlation screen was carried out. Using CWP as an exposure factor, we had to modify the genome-wide relevance criterion (*p* < 5×10^-6)^ to acquire more meaningful SNPs (17). For the purpose of the reverse MR analysis with GERD as a trigger factor, we only included SNPs with a significant connection and genome-wide significance level (*p* < 5×10^-8^). To reduce the impact of severe linkage disequilibrium, we employed tight selection criteria for SNPs, such as R^2^ = 0.001, with a genetic frame of 10,000 kb in the European 1000 Chromosome standard panel (18). We utilized PhenoScanner V2 (http://www.phenoscanner.medschl.cam.ac.uk) to determine whether the SNPs employed were associated with important potential confounding factors (19), and if so, excluded them. We evaluated the statistical significance of each SNP using the *F* -value (*F* = *β*^2^/SE^2^), where *β* describes the allele effect value and SE denotes the standard error. Excluded SNPs having *F*-value <10 (20). Finally, we identified genuine SNPs that are substantially related with GERD or CWP.

### Sensitivity analyses

The Cochran’s Q test allowed us to determine the presence of heterogeneity among those risk factors. A *p* -value ≥ 0.05 suggests a low chance of heterogeneity (21). To see if other factors were influencing the outcomes, we employed the MR Egger’s intercept test (22). The occurrence of substantial disparities highlighted the necessity to explore other variables. To locate any outliers that might affect the results, the MR pleiotropy residual sum and outlier (MR-PRESSO) test was applied (23).

We reanalyzed the data after subtracting the outliers to eliminate any other factors that could have biased the results. To ascertain the impact of genetic variation, we also implemented a leave-one-out sensitivity analysis (24).

### MR analysis

Inverse variance weighted (IVW) analysis was the main statistical method used in this work (24). In parallel, stability analyses and findings validations were carried out using the weighted median and MR-Egger regression models (22, 25). The study yielded odds ratios (OR) and 95% confidence intervals (95% CI) for the MR results.

### Statistical methods

Version 5.1.0 of the R software (26) ’s “TwoSample MR” package was used for all statistical studies. *p* < 0.05 was used for demonstrating statistical significance.

## Results

### Genetic instrumental variables

Following a rigorous surveillance, we narrowed our selection to 7 separate SNPs for GERD and 28 SNPs for CWP. To do this, we eliminated chain imbalances and associated confounders, with smoking being recognized as a risk factor for GERD and smoking, obesity, mental illness, cancer, and osteoarthritis as risk factors for CWP (27–29). Furthermore, SNPs that did not appear in the endpoint GWAS were deleted. Following this, it came to light that all SNPs had *F* statistics > 10, demonstrating that the causal conclusions obtained in our investigation can be understood without taking into account weak SNPs. S1 Table summarizes our findings and offers a detailed overview of the combined information from the identified SNPs.

### Causal effect of CWP on the risk of GERD

To determine the robustness of the causal link between GERD and CWP, a sensitivity analysis was performed. Heterogeneity among SNPs was demonstrated by the Cochran’s Q *p*-values in MR-Egger (Q = 38.443, *p* <0.001) and IVW (Fig 2, Q = 40.197, *p* < 0.001). As an outlier, rs1491985 was removed based on the initial MR PRESSO result (Table 1, *p* = 0.0001<0.05). rs923593 and rs7541613 were eliminated as well since the second MR PRESSO result suggested they might be potential outliers (Table 1, *p* = 0.018<0.05). There were no outliers in the third MR PRESSO data (Table 1, *p* = 0.076 > 0.05). For the final MR analysis, 4 SNPs were chosen based on the analysis above. Even with the continuous heterogeneity [MR-Egger (Q=10.116, *p* = 0.006) and IVW (Fig 2, Q=12.035, *p* = 0.007)], the accuracy of the MR results would remain unaffected using the random-effects IVW model. This study did not exhibit pleiotropy, according to the MR-Egger intercept test (Fig 2, *p* = 0.601). Additionally, a leave-one-out analysis was carried out, eliminating SNPs one at a time, and it was shown that no SNP had a statistically significant impact on the entire results (Fig 3A).

**Fig 2.**
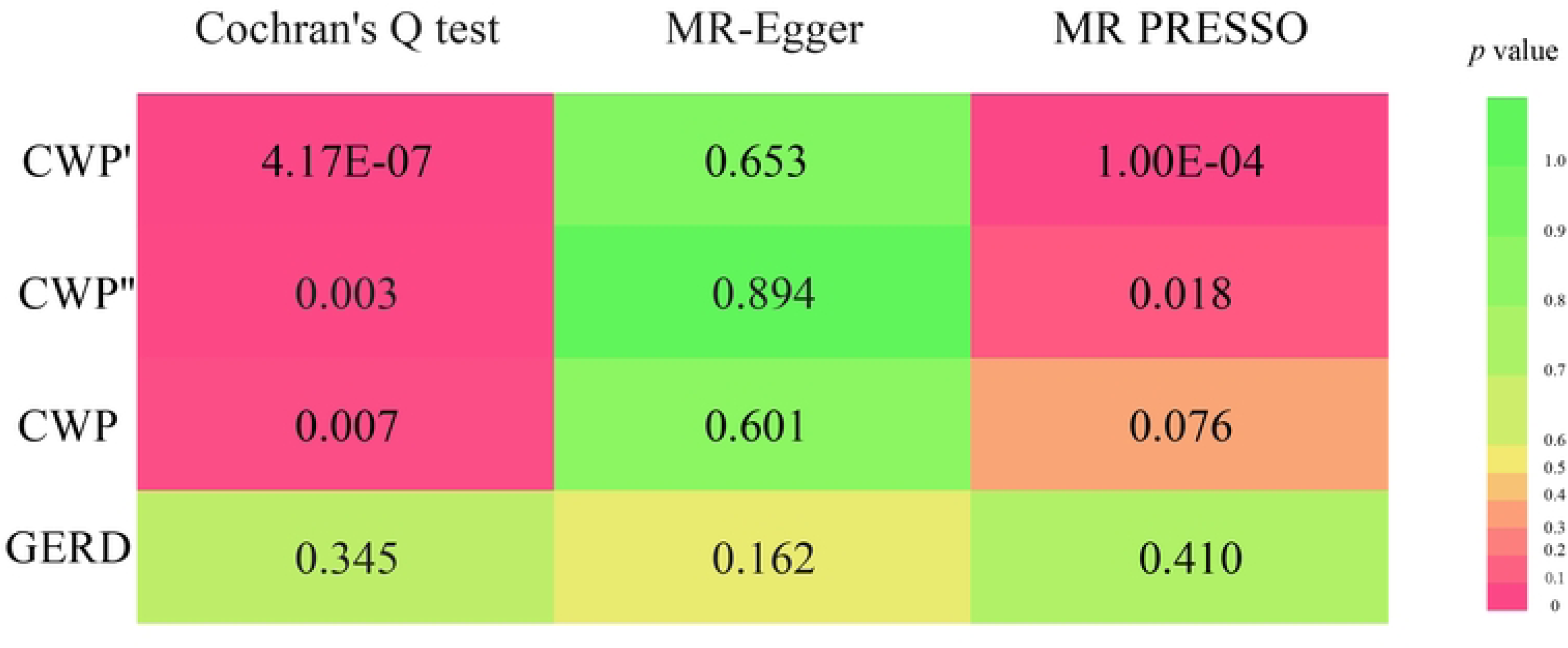
Results of sensitivity analysis. (CWP’: SNPs of CWP used in the first Mendelian analysis; CWP’’: SNPs for CWP used in the second Mendelian analysis, CWP: SNPs for CWP used in Mendelian analysis after removing all outliers.)

**Fig 3.**
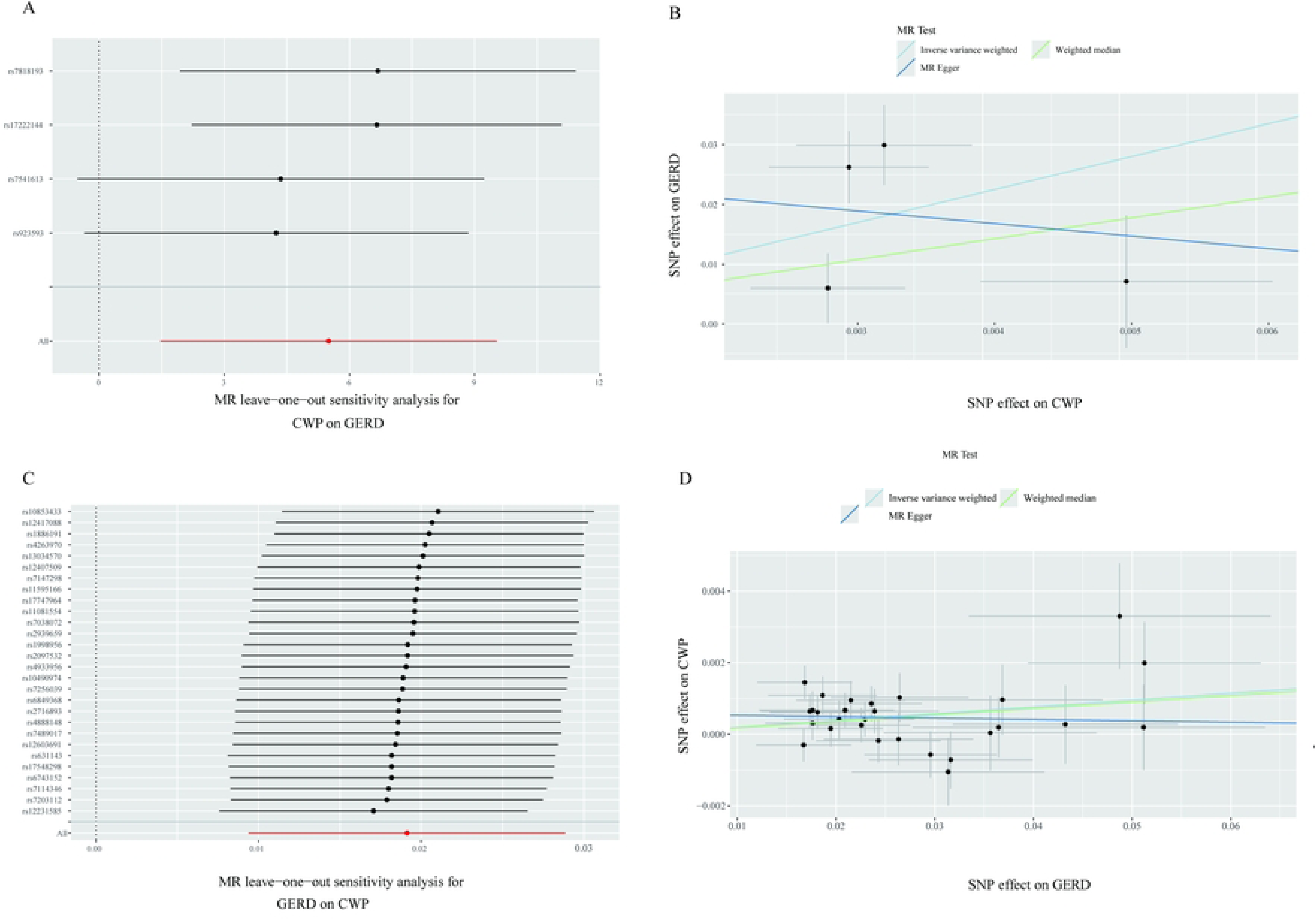
Results of MR analysis. (A-Scatter plots of estimates for the association of GERD on CWP; B-Leave-one-out results for GERD on CWP; C-Scatter plots of estimates for the association of CWP on GERD; D-Leave-one-out results for CWP on GERD)

**Table 1.** Results of MR analysis.

The results of IVW showed that the genetically predicted prevalent population would have a significantly increased risk of GERD compared to those without CWP (N_SNP_ = 4, OR: 245.244; 95% CI: 4.35E+00,1.38E+04; *p* =0.007 < 0.05) (Table 1). The result of the weighted median method also corroborated the trend observed in the IVW analysis. Nevertheless, the MR-Egger results weren’t compatible with the IVW direction. The two methods did not yield statistically significant results (Table 1).

To determine how the outliers affected the final MR results, MR analysis was also done for each SNP before the outliers were eliminated. The IVW results demonstrated that the MR data support the function of CWP in raising the chance of suffering from GERD, even with outliers (S2 Table, S1 Fig). These findings indicate that CWP patients is a causal risk factor for GERD.

### Causal effects of GERD on the risk of CWP

Similarly, sensitivity studies were carried out to determine the strength of the causal link between GERD and CWP. There was no indication of possible directed pleiotropy according to the results of the MR-Egger intercept test (Fig 2, *p* = 0.162) or MR PRESSO findings. The Cochran’s Q p-values in the IVW (Figure 2, Q = 27.171, *p* = 0.400) and MR-Egger (Q = 29.339, *p* = 0.345) approaches indicated a low chance of heterogeneity. MR PRESSO was performed, and there were no significant outliers (Fig 2, *p* = 0.41), indicating that the MR data were credible. Furthermore, leave-one-out analysis revealed no significant differences in the associations found when any one SNP was deleted (Fig 3C).

Our results offer strong evidence that GERD and CWP are causally related. The study discovered that people with GERD have a greater chance of having CWP (N_SNP_ = 28; OR:1.019; 95% CI: 1.009-1.029; *p* = 0.029 < 0.05) (Table 1). The weighted median technique confirms the trend found in the IVW analysis (N_SNP_ = 28; OR: 1.018; 95% CI: 1.004-1.033; *p* = 0.013<0.05) (Table 1). Nonetheless, the MR-Egger results did not match the IVW orientation (Fig 3D).

## Discussion

Our findings provide solid proof that GERD and CWP are causally connected. Individuals with a genetic predisposition to GERD may be at a higher risk for CWP. Those with CWP have a higher chance of acquiring GERD. We verified these findings with a sensitivity analysis, confirming causality between these two diseases. Several hypotheses try to explain the link between CWP and GERD. The potential causes include psychosocial variables as well as brain-gut interactions.

Psychosocial factors such as negative childhood memories, catastrophic life events, and interpersonal disputes all have a substantial impact on the onset, severity, and susceptibility of CWP (30). Prolonged psychological discomfort can lead to CWP in persons with psychological illnesses such as anxiety and depression (31). Gastric reflux into the throat can be caused by mood disorders by psychological stress that restricts the lower part of the sphincter. Furthermore, it can increase the gastrointestinal tract’s sensitivity to stomach acid (32). Additionally, numerous stressful situations and psychiatric illnesses might exacerbate the symptoms of GERD (33).

The gut-brain axis and the hypothalamic-pituitary-adrenal (HPA) axis are hypothesized to play roles in the pathophysiology of certain disorders. When the brain detects stress, the HPA axis is engaged, and the hypothalamus produces corticotropin-releasing factor (CRF), which is found in both the brain and the gut (8). Overproduction of CRF can impair intestinal motility, causing greater pain and visceral sensitivity. Stressful life events can permanently overproduce CRF and affect the sympathetic and HPA axes (34, 35). The gut-brain axis is regulated bidirectionally. The central nervous system (CNS) can control intestinal pain symptoms and reduce hypersensitivity. The gut flora can interact with the CNS via influencing the levels of gut neurotransmitters and cytokines, either directly or indirectly. The association between FM and gut bacteria has been proven, with *Coprococcus2*, *Eggerthella*, and *Lactobacillus* being reported to increase the risk of FM (36).

Risk factors for GERD and CWP may be similar, including depression, smoking, and sleep disturbances (29, 37). To prevent inclusion bias from observational studies that neglect to omit mutual factors, relevant confounders were eliminated before to the two-sample MR analysis in the current investigation, and GERD and CWP were included as distinct outcomes. The degree of evidence from observational studies and the superiority of MR analysis (38) meant that confounders had less of an impact on the current study’s conclusions. Despite the fact that there is heterogeneity among the instrumental factors for positive MR, IVW’s random effects model ensures that the results are reliable despite the heterogeneity.

Although the screening thresholds were relaxed, only a small number of genome-wide significant SNPs were found in the CWP GWAS, which may explain the large variability in point estimates across MR methods. This could account for the considerable variation in point estimates amongst MR techniques (39). Furthermore, fibromyalgia and/or chronic localized musculoskeletal pain are included in the broad diagnosis of CWP. Inaccurate definitions have the potential to add confounding variables to unprocessed GWAS data, decreasing their statistical power. Integrating the distinct GERD and CWP phenotypes is a crucial topic of investigation for subsequent investigations. On the one hand, since the research solely employed GWAS data from European groups, it avoids the influence of other populations. It does, however, restricts how broadly the findings may be extended to various populations.

In the future, more research with datasets from different populations will be needed. Ultimately, this study deduced the genetic basis of the causal association between GERD and CWP. Nevertheless, the underlying biological mechanisms remain incompletely understood, and further studies are essential to validate this assertion in the future.

## Conclusion

Our findings indicate that GERD and CWP have a bidirectional causal connection, meaning that having one illness raises the chance of getting the other. These findings support earlier studies and point to the necessity of treatments that can deal with both diseases simultaneously.

## Data Availability

If the data are all contained within the manuscript and/or Supporting Information files.

http://www.phenoscanner.medschl.cam.ac.uk

## Acknowledgments

Every author—MC, HT, JZ, YZ, SW, YX, and LH—contributed significantly to the study’s execution. Their main contributions are as follows: (1) they planned and created the research; (2) they drafted and edited the article; and (3) they submitted the final manuscript for approval. They also collected, analyzed, and interpreted data. Simultaneously, MC assumed control of the entire study procedure.

We appreciate the investigators and subjects of the initial MR investigation. We acknowledge the provision of this data by IEU GWAS and the UK Biobank. Author contributions

## Supporting information

**S1 Fig. Results of MR analysis before excluding outliers.** (A-Scatter plots of estimates for the association of GERD on CWP’; B- Leave-one-out results for GERD on CWP’; C- Funnel plots of estimates for the association of GERD on CWP’;D- Scatter plots of estimates for the association of GERD on CWP’’; E- Leave-one-out results for GERD on CWP’’; F- Funnel plots of estimates for the association of GERD on CWP’’.)

**S1 Table. SNPs used as genetic instrumental variables for GERD or CWP.** (EAF-effect allele frequency, SE-standard error, CWP’- SNPs of CWP used in the first Mendelian analysis, CWP’’- SNPs for CWP used in the second Mendelian analysis, CWP- SNPs for CWP used in Mendelian analysis after removing all outliers.)

**S2 Table. Results of MR analysis before excluding outliers.**

